# Perspectives of Heart Transplant Patients and Providers on Acute Rejection Surveillance: A Mixed-Methods Study

**DOI:** 10.1101/2024.12.29.24319749

**Authors:** Hyoungmin Kim, Vincenzo Cusi, Melissa McLenon, Jose Benjamin Cruz Rodriguez, Quan M. Bui, Jennifer Chak, Marcus Anthony Urey, Justin Cole, Rebecca Fielding-Miller, Paul J. Kim

## Abstract

**Background:** Endomyocardial biopsies (EMB) remain the reference standard for detection of acute rejection in heart transplant (HTx) patients. Recent studies evaluating novel noninvasive tests have sparked a renewed discussion in the HTx community about revising acute rejection surveillance policies. However, patient and provider perspectives remain underexplored. This single-center study examined both HTx patient and provider perspectives on replacing EMBs earlier with noninvasive blood tests.

**Methods:** We performed semi-structured interviews with 28 HTx patients to explore their perspectives on replacement of EMBs with donor-derived cell-free DNA (dd-cfDNA) early post-HTx. We subsequently conducted a survey of 118 HTx patients using self-administered online questionnaires. We also performed semi-structured interviews with 18 HTx providers to explore their perspectives. Thematic analysis was performed on interview and open-ended survey responses using deductive and inductive approaches. Patient quantitative survey responses were analyzed with descriptive statistics.

**Results:** Our study identified three key themes: patient anxiety related to EMBs, importance of patient-provider communication, and strong interpersonal trust in providers by HTx patients. Although 78.4% of patients experienced EMB-related anxiety, they prioritized testing accuracy to ensure “the health of their new heart.” Consequently, patients favored the most accurate testing protocol and trusted providers to make this decision (91.1%). HTx providers raised concerns about the accuracy and safety of noninvasive surveillance testing for high risk patients.

**Conclusion:** HTx patients trusted their providers to determine the most accurate acute rejection surveillance policy. Additionally, our study provides important patient-centered priorities to guide the implementation of early noninvasive testing into clinical practice.

## INTRODUCTION

While EMBs are the reference standard of detecting acute rejection post-HTx, there is active research to replace surveillance EMBs earlier with noninvasive testing, due to potential complications associated with EMBs.^1–3^ The IMAGE trial was the first randomized controlled trial (RCT) that demonstrated the safety of replacing surveillance EMBs with gene-expression profiling (GEP) blood tests in low risk HTx patients.^4^ More recently, novel noninvasive tests, such as dd-cfDNA testing, have shown promise in further reduction of surveillance EMBs in the early post-HTx period and also in detecting antibody-mediated rejection.^3,5,6^ As a result, a number of HTx centers have changed their institutional policies to increasingly utilize noninvasive tests in place of surveillance EMBs, starting as early as 2 to 4 weeks post-HTx.^1,5,7^ However, the perspectives of HTx patients and providers on proposed changes to acute rejection surveillance policy have not been fully explored.

This study leveraged a unique opportunity to obtain patients’ and providers’ perspectives on EMBs at our institution, where policy changes to replace EMBs with dd-cfDNA are being considered early post-HTx. We hypothesized that the majority of HTx patients would strongly advocate for replacing EMBs earlier with noninvasive testing. These findings will inform the integration of early noninvasive testing into future patient-centered policy and guideline changes.^8^

## MATERIALS & METHODS

### Study Design

This was a mixed-methods study conducted at University of California, San Diego (UCSD) of HTx patients who underwent EMB and the HTx providers who treat these patients. This study was performed in 2 parts: a qualitative part of semi-structured interviews of HTx patients and providers and a quantitative part of a larger cohort of HTx patients using a self-administered online survey. This study was overseen by a multidisciplinary research team with expertise in HTx (VC, MM, JBCR, QMB, MAU, and PJK) and qualitative methodology (QMB and RFM). We followed the Consolidated Criteria for Reporting Qualitative Research (COREQ) and Standards for Reporting Qualitative Research (SRQR) guidelines to report this study.^9,10^

### Participants

Adult HTx patients, who were 18 years of age or older, transplanted from August 2021 to June 2023, and had previously undergone at least one EMB, were recruited consecutively during scheduled clinic visits for semi-structured interviews (conducted from June to August 2023). At our institution, surveillance EMBs range from 4-10 months post-HTx, dependent on HTx patient risk (**Appendix A**). We subsequently invited HTx patients transplanted from August 2019 to July 2023 to take part in an online survey that also allowed for open-ended responses.^1^ We also conducted semi-structured interviews of HTx providers from January to March 2024. HTx providers included board-certified HTx cardiologists, advanced practice providers, nurses, or radiologic technologists who were 21 years of age or older, had been practicing for at least 3 years, and directly cared for HTx patients.

### Data Collection

Interview guides were developed for HTx patients (**Appendix B**) and providers (**Appendix C**) based on discussions with the core research team (HK, VC, MM, QMB, MAU, RFM, and PJK) and were iteratively refined. Firstly, a semi-structured interview guide was developed that explored patients’ perspectives on replacing EMBs earlier with noninvasive blood tests. Subsequently, a semi-structured interview guide was developed for HTx providers. Consistent with qualitative methodology, the interview guide was modified during the study as new topics emerged from the interviews.^11,12^ We then used preliminary results from the qualitative analysis of patient responses to develop a brief self-administered online survey distributed using Qualtrics software (Qualtrics International Inc., Seattle, Washington, USA; **Appendix D**). The survey was available to patients in English and Spanish–the two most common languages at UCSD. The survey was not pilot tested prior to its distribution. The online survey responses were anonymized to allow for critical feedback from patients and for HIPAA compliance per UC San Diego Office of IRB Administration policy.

### Analysis

Interviews were conducted and recorded by HK, VC, and MM. We utilized a standardized telephone script prior to interviewing semi-structured interview patients and providers. Recorded interviews were transcribed using Otter.ai (Otter.ai Inc., Mountain View, California, USA) and were verified for accuracy by HK and VC. Transcripts were not returned to participants for comments or corrections. Transcripts were coded and thematic analyses were performed by HK, VC, and PJK. A hybrid thematic process combining inductive and deductive approaches was followed, grounded in a post-positivist paradigm.^13^ Initial codes were developed deductively based on prior clinical and research experience related to EMBs. The codes reflected concepts such as “anxiety,” “pain,” “rejection surveillance,” “communication of findings,” “reduction of frequency of EMBs,” and more. However, our analysis remained open to inductive insights from participant narratives. Interviews continued until code and meaning saturation were reached, supervised by PJK, QMB, and RFM. Confidentiality of data was kept by deidentifying patient and provider responses and assigning unique code numbers. Qualitative data was managed using Microsoft Excel 16.1 (Microsoft Corporation, Redmond, Washington, USA). Qualitative analysis of patient interviews was used to guide quantitative analysis of the online survey as is standard in mixed sequential designs.^14^ Quantitative online survey data was exported and analyzed with descriptive statistics using R (R Core Team, 2025). Fisher’s exact test was used to compare patient satisfaction scores using patient experience surveys from NRC (National Research Corporation, Lincoln, Nebraska, USA) Health that were ≥ 9 from a scale from 0-10 (0 being not at all satisfied and 10 being extremely satisfied) 6 months before and after implementation of suggestions by HTx patients and providers to improve the patient EMB experience. Thematic analysis was also performed for the open-ended responses from the online survey.

### Regulatory Ethics

All invited survey patient participants were eligible to participate in a drawing to win one of ten $50 gift cards. Ethics approval was granted by the UCSD Office of IRB Administration (No. 805675). This study adheres to the principles of the Declaration of Helsinki formulated by the World Medical Association and the US Federal Policy for the Protection of Human Subjects. All interviewed subjects gave their informed consent prior to their inclusion in the study.

## RESULTS

### Respondent characteristics

For the semi-structured patient interviews, 56 participants were invited and 28 (50.0%) agreed to participate. For the online survey, 286 patient participants were invited and 118 surveys were completed (41.3%). HTx provider interviews included 7 HTx physicians, 5 registered nurses (RN), 4 advanced practice providers (APP), and 2 radiologic technologists (RT).

Respondent characteristics for interviewed patients are shown in **Table 1**. The number of sensitized patients (panel of reactive antibodies ≥ 10%) was 14.3%.^3^ There were 18 (64.3%) low, 4 (14.3%) moderate, and 6 (21.4%) high risk patients. All EMBs were performed using fluoroscopy guidance and 86.7% were performed using the right internal jugular vein as access.^15^ The median number of EMB per patient participant at the time of the study was 8 (IQR, 7 - 10). Most EMBs were performed in an outpatient setting (76.4%) and performed for surveillance indication (83.5%). The median time from HTx to semi-structured interviews was 89 days (IQR, 52 - 172 days). Interviews lasted a median time of 14 minutes (IQR, 12 - 17.5 minutes).

**Table 1.** Patient Characteristics for Semi-Structured Interviews (n = 28 patient participants).

| **Table 1.** |  |
| --- | --- |
| **Recipient characteristics** |  |
| Age, y, mean (SD) | 60.1 (10.4) |
| Male, N (%) | 22 (78.6) |
| Race |  |
| Asian, N (%) | 1 (3.6) |
| Black, N (%) | 2 (7.1) |
| Other Race, N (%) | 6 (21.4) |
| Pacific Islander, N (%) | 1 (3.6) |
| White, N (%) | 18 (64.3) |
| Ethnicity |  |
| Hispanic or Latino, N (%) | 3 (10.7) |
| Recipient BMI, mean (SD) | 27.2 (6.6) |
| Indication for Transplant |  |
| NICM, N (%) | 16 (57.1) |
| ICM, N (%) | 11 (39.3) |
| Re-HTx, N (%) | 1 (3.6) |
| Allosensitization pre-HTx<br>(PRA ≥ 10%), N (%) | 4 (14.3) |
| Patient Risk Status |  |
| Low Risk (%) | 18 (64.3) |
| Moderate Risk (%) | 4 (14.3) |
| High Risk (%) | 6 (21.4) |
| Durable MCS at time of HTx,<br>N (%) | 6 (21.4) |
| **HTx characteristics** |  |
| Multiorgan transplant, N (%) | 4 (14.3) |
| Induction therapy, N (%) | 9 (32.1) |
| De novo DSA, N (%) | 5 (17.9) |

Abbreviations: BMI, body mass index; DSA, donor-specific antibody; HTx, heart transplantation; ICM, ischemic cardiomyopathy; MCS, mechanical circulatory support; NICM, nonischemic cardiomyopathy; PRA, panel reactive antibodies

### HTx patient perspectives

#### Semi-structured interviews

Three common themes were identified: (1) patient anxiety related to EMBs, (2) importance of patient-provider communication, and (3) strong interpersonal patient trust in providers by HTx patients (**Table 2**).

**Table 2.** Major Themes, Representative Quotations, and Proposed Approaches to Improve the EMB Patient Experience.

| Major Themes | Illustrative Quotations | Proposed Approaches |
| --- | --- | --- |
| Patient Anxiety | *"If they take a little longer after the anesthesia kicks in, [the biopsy experience] will be 100% better. Right now [it] is 95%."*<br>(Survey Participant 24) | • Allow time for lidocaine to settle.<br>• Communicate waiting room delays promptly.<br>• Ask for patient's preferences in music during the EMB |
| Patient-Provider Communication | *"Every time they ask me how are you feeling, how are you doing. They ask you about 10 times during the biopsy. That's what makes me feel good."*<br>(Patient Participant 7) | • Introduce patients to the HTx team prior to draping.<br>• Check-in with the patient regularly.<br>• Provide a simple explanation of EMB results in a timely fashion. |
| Trust in Providers | *"The doctors explained it so many times what they're doing and why they're doing it [that] I trust what they're gonna do."*<br>(Patient Participant 22) | • Acknowledge patient anxiety related to EMBs.<br>• Address communication barriers throughout the EMB process.<br>• Include patient participation in their own care before, during, and after EMBs. |

#### Patient anxiety related to EMBs

Interviewed patients commonly described anxiety related to EMBs. This anxiety often started in the waiting room for patients and was exacerbated by delays that were not communicated to patients. In addition, a few patients described increased pre-procedural anxiety due to prior negative EMB experiences.

> *‘I [get] anxious a little bit because of that initial time when I was like tremoring on the table. I’m always afraid it’s gonna hurt really bad… it was just like, horrible pain. And you know, because of that I do get anxious before the procedures.’ – Interview Patient Participant 2*

> *‘My apprehension and anxiousness increases when there is a delay… I have to sit there and never know exactly when I’ll be taken… it’s just not a good experience.’ – Interview Patient Participant 21*

Patients frequently cited that the most distressing and painful part of EMBs was the lidocaine injection. Suggestions for changes to the EMB experience to reduce anxiety were also provided by patients (**Table 2**).

> *‘If they take a little longer after the anesthesia kicks in, [the biopsy experience] will be 100% better. Right now [it] is 95%.’ – Survey Participant 24*

> *‘I just want to be able to look around and see the room and not feel so confined underneath the whole cover itself.’ – Interview Patient Participant 1*

> *‘[I told the fellow who did my biopsy] for the future [to] come around and introduce yourself because that would have been nice to know.’ – Interview Patient Participant 27*

While a few HTx patients may experience negative EMB experiences, many interviewed patients considered surveillance EMBs a necessity despite the potential pain associated with EMBs.

> *‘I think it’s vitally important that if there is something going on that they get ahead of it early on, [catch it], [and] get rid of it.’ – Interview Patient Participant 1*

> *‘Even if everything was really painful, it’s totally worth it. You’re not gonna hear complaints from me.’ – Interview Patient Participant 12*

A minority (32.1%) of interviewed patients expressed their desire for alternatives to frequent surveillance EMBs due to their anxiety, previous poor EMB experience, and perceived risks. Additionally, some of the perceived EMB risks were attributed to information from sources other than the patients’ providers.

> *‘I think [the EMB frequency] is a little bit too much. Now, it’s like every two weeks but it will [get] better. It will be like one in a month and then one in a year. [Right] now it’s just the beginning, so it’s hard.’ – Interview Patient Participant 15*

> *‘A yearly biopsy where there are no signs of rejection or cause for concern seems unnecessary considering the risk [of EMBs].’ – Survey Participant 15*

> *‘[I would prefer less biopsies] because my wife said that [EMBs] are dangerous.’ – Interview Patient Participant 6*

#### Importance of patient-provider communication

Compassionate and consistent patient-provider communication was considered an important component by patients before, during, and after EMBs (**Figure 1** and **Table 2**). Good patient-provider communication not only reduced patient anxiety but also allowed patients to feel included in their care.

> *‘You have a heart transplant and every doctor who comes in knows your whole life story… We went through everything and asked me questions, they already knew where I grew up, where I’m from, and that I have kids and stuff like that… They knew [about me] and that helped a lot.’ – Interview Patient Participant 14*

**Figure 1.**
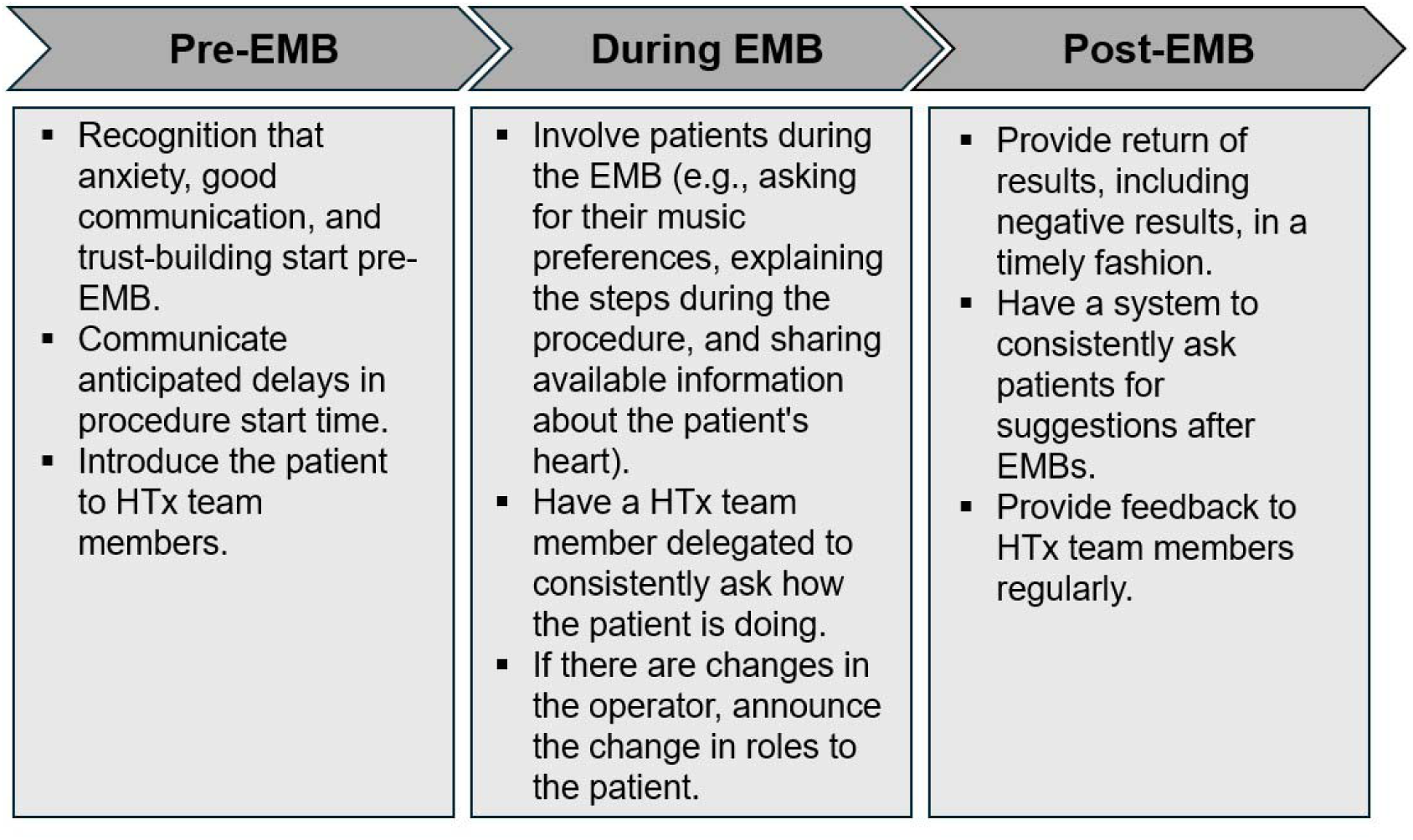
Anxiety, Communication, and Trust-building throughout the patient journey for EMBs. Patients and providers provided practical suggestions to reduce patient anxiety, improve patient-provider communication, and strengthen patient trust throughout the three stages of a patient’s EMB experience (before, during, and after).

> *‘It helps to calm the patient or make them feel more comfortable if they’re not left laying on the table by themselves staring at the wall while the staff is on the other side of the room talking.’ – Survey Participant 109*

> *‘[The] last time I went, I got to see everything on the screen. And [the doctor] was explaining to me what those things were [on] the sternum.. I [saw] the little wings and [the doctor said] ‘oh, you got extra hardware in there’… I learned about that.’ – Interview Patient Participant 6*

Patients also stated their desire for the return of their biopsy results in a timely manner and in a language understandable to them.

> *‘But if you don’t hear from the team, it’s just assumed that the biopsy was either a zero or a one… I would prefer to see that as a last result in my medical records, so I don’t have to wonder.’ – Interview Patient Participant 21*

> *‘I’d like to look at pictures… [the EMB results] doesn’t really tell me much [as a layman].’ – Interview Patient Participant 2*

#### Strong interpersonal trust in providers by HTx patients

HTx patients commonly described their strong interpersonal trust with their providers. A subtheme identified was that patients felt the final decision related to acute rejection surveillance should ultimately come from their HTx team. Patients who had negative experiences with EMBs also continued to trust their providers’ recommendations.

> *‘I’ve never had a heart transplant before this one. I don’t have a professional opinion on that. I just know what the doctors are saying [and] this is what we need to do.’ –*

#### Interview Patient Participant 1

> *‘I’ve got scar tissue on my neck now… you know, it hurts… but [the doctors] know what they’re doing… so I comply.’ – Interview Patient Participant 2*

> *‘I think [determining biopsy frequency] should really be up to the cardiologists… The end goal surpasses any kind of inconvenience.’ – Interview Patient Participant 12*

Interpersonal trust was established at the beginning of the relationship with HTx providers and patients. This trust was strengthened further through consistent and compassionate communication, building a sense of camaraderie with the HTx team.

> *‘I put my hands up to the doctor six years ago now. I said I tried doing it myself [and] it didn’t work… the team has been awesome.’ – Interview Patient Participant 17*

> *‘One thing you guys did [that] was really good was introduce the entire cardiology team on the front end. I [had] nice conversations with everybody. It’s kind of like a camaraderie.’ – Interview Patient Participant 12*

> *‘The doctors explained it so many times what they’re doing and why they’re doing it [that] I trust what they’re gonna do.’ – Interview Patient Participant 22*

Gratitude and a feeling of indebtedness were also frequently associated with strong interpersonal trust in the HTx team.

> *‘I got a new lease on life. The heart they pulled out of me was down to 8%. So for me, I have two birthdays… I have to do whatever they want… Just trying to comply to the best of my ability so that I can be well.’ – Interview Patient Participant 2*

> *‘If I had to do it over again. I would do exactly what you guys said because the way you guys took care of me is undescribable [sic].’ – Interview Patient Participant 25*

Ultimately, patients wanted the assessment of their new heart’s condition to be accurate, regardless of the surveillance strategy used. Patients would consider alternatives if their HTx team considered them to be as accurate as the current acute rejection surveillance policy.

> *‘[Changes to patients’ EMB policy] should really be up to the cardiologists who live with their decisions… if they do see a significant amount of people losing their lives, what is that worth? Even if you lose one I [think decreasing EMBs] is not worth it.’ – Interview Patient Participant 12*

> ‘You guys know what you’re doing. So if you guys think you can cut the number of times that you can do the biopsy with success, I would say that’s great. But, if there’s a *problem [with doing that, then] keep going the way you’re going.’ – Interview Patient Participant 25*

> ’If it was determined that the outcomes are the same, that would be great… If [EMBs] were less frequent and have the same outcomes… I can’t imagine anybody would be against that.’ – Interview Patient Participant 23

### Quantitative (patient online survey questionnaire)

Surveyed patients recalled their average pain associated with EMBs low (2.5 out of 10; **Table 3**). In addition, anxiety associated with EMBs decreased with subsequent EMBs in the majority (78.4%) of survey participants.

**Table 3.**
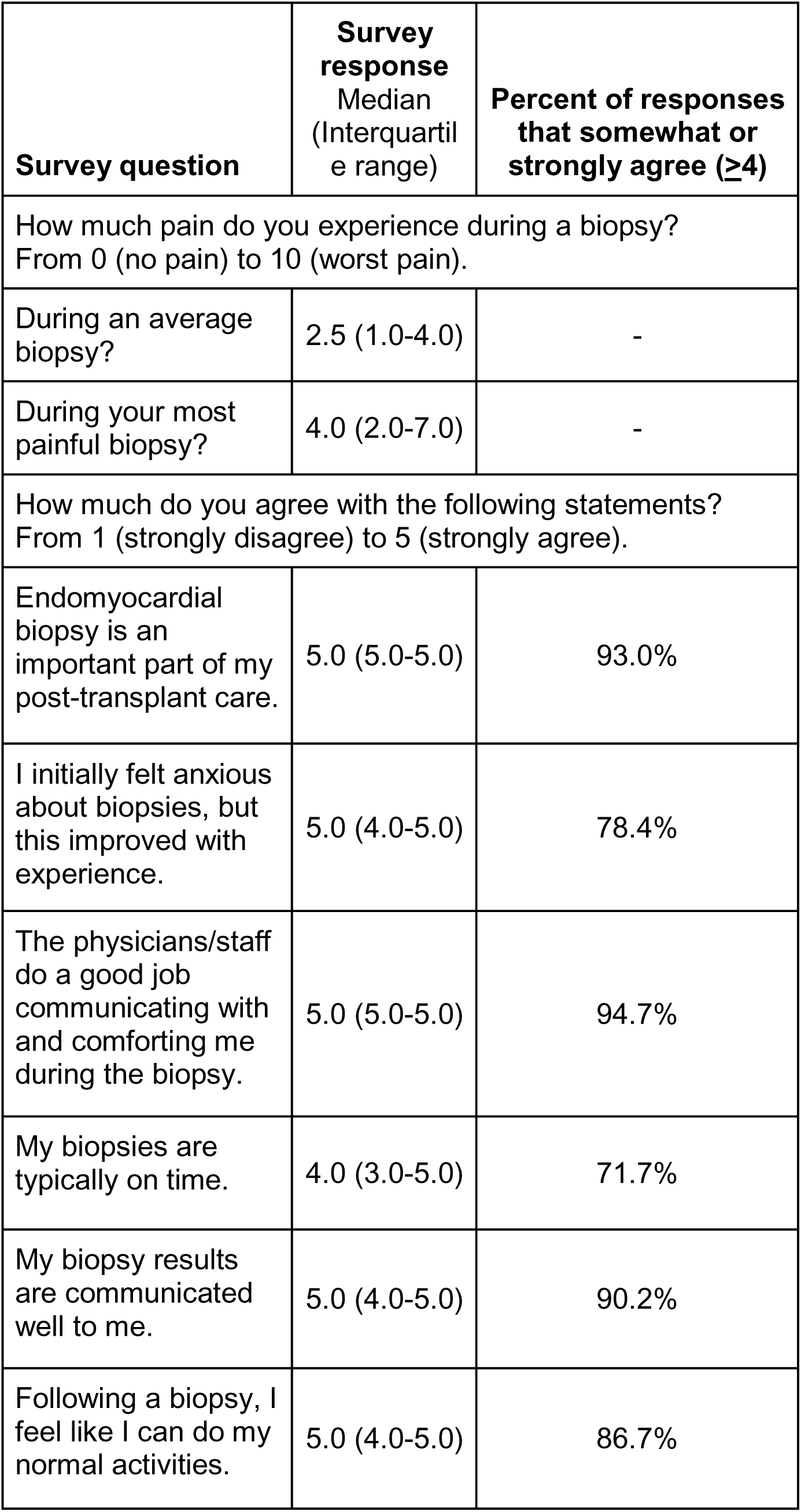
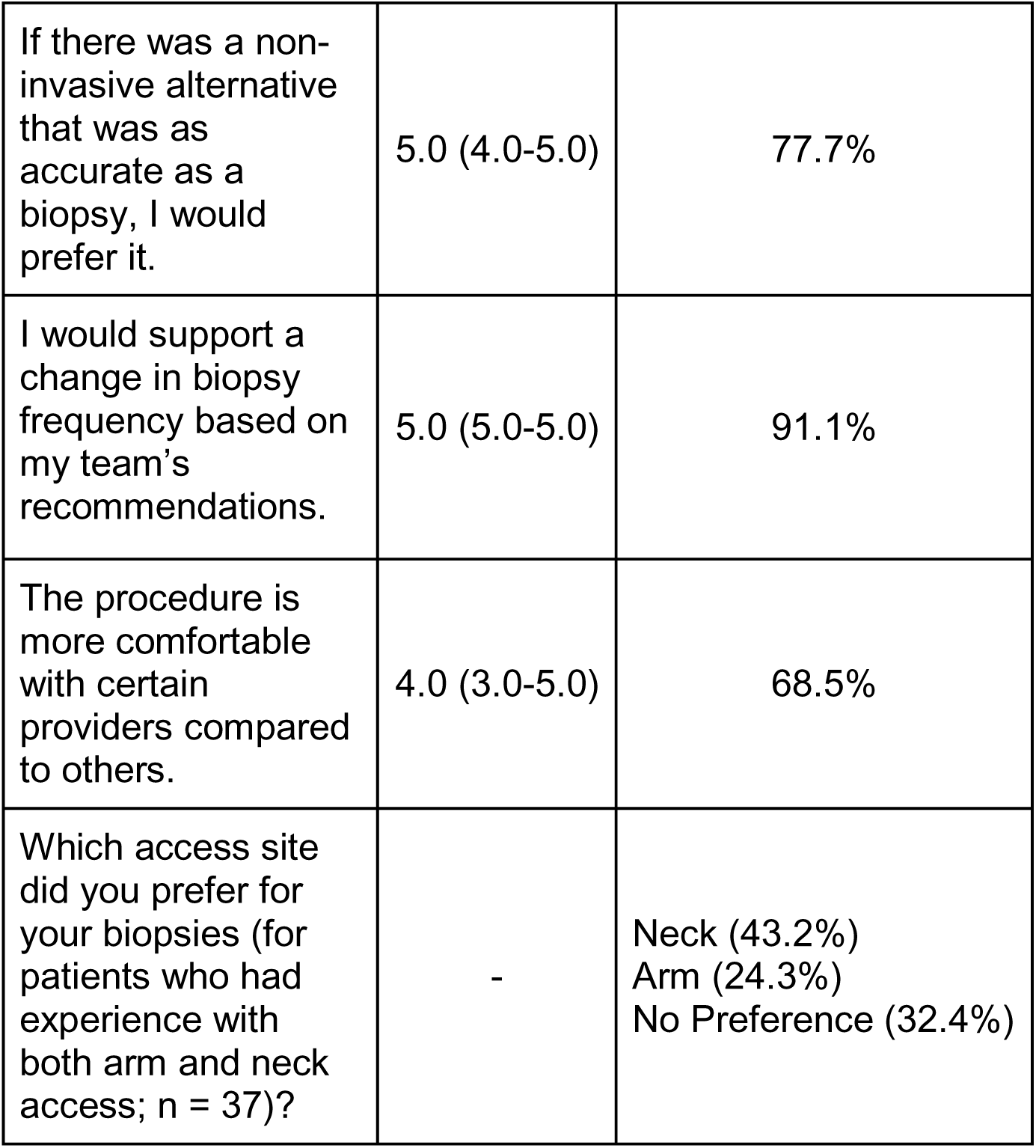
Patient Online Survey Results (n = 118 patient participants).

| **Survey question** | **Survey response**<br>Median<br>(Interquartile range) | **Percent of responses that somewhat or strongly agree ($\geq 4$)** |
| --- | --- | --- |
| How much pain do you experience during a biopsy?<br>From 0 (no pain) to 10 (worst pain). |  |  |
| During an average biopsy? | 2.5 (1.0-4.0) | - |
| During your most painful biopsy? | 4.0 (2.0-7.0) | - |
| How much do you agree with the following statements?<br>From 1 (strongly disagree) to 5 (strongly agree). |  |  |
| Endomyocardial biopsy is an important part of my post-transplant care. | 5.0 (5.0-5.0) | 93.0% |
| I initially felt anxious about biopsies, but this improved with experience. | 5.0 (4.0-5.0) | 78.4% |
| The physicians/staff do a good job communicating with and comforting me during the biopsy. | 5.0 (5.0-5.0) | 94.7% |
| My biopsies are typically on time. | 4.0 (3.0-5.0) | 71.7% |
| My biopsy results are communicated well to me. | 5.0 (4.0-5.0) | 90.2% |
| Following a biopsy, I feel like I can do my normal activities. | 5.0 (4.0-5.0) | 86.7% |
| If there was a non-invasive alternative that was as accurate as a biopsy, I would prefer it. | 5.0 (4.0-5.0) | 77.7% |
| I would support a change in biopsy frequency based on my team's recommendations. | 5.0 (5.0-5.0) | 91.1% |
| The procedure is more comfortable with certain providers compared to others. | 4.0 (3.0-5.0) | 68.5% |
| Which access site did you prefer for your biopsies (for patients who had experience with both arm and neck access; n = 37)? | - | Neck (43.2%)<br>Arm (24.3%)<br>No Preference (32.4%) |

Most patients indicated that EMBs are an important part of their post-HTx care (93.0%) and would support a change in EMB frequency based on their HTx team’s recommendations (91.1%). Additionally, if noninvasive testing is as accurate as surveillance EMBs, 77.7% of surveyed patients would prefer noninvasive testing over surveillance EMBs. The patients that continued to prefer surveillance EMBs over noninvasive testing most often cited their gratitude in their HTx team in their open-ended responses. These results supported our qualitative findings.

> *‘As a whole, you all do a great job of making me feel comfortable through the procedure. Thank you all very much.’ – Survey Participant 45*

> *‘I’m always very appreciative of the high level of care the Docs and staff in the Cath lab give me. After about 20 - 25 biopsies many of these people have treated me multiple times. Thanks to them all.’ – Survey Participant 63*

### HTx Provider Perspectives

The majority (87.5%) of HTx physicians perceived positive patient experience with EMBs. This perception was reduced in HTx non-physician providers (APPs, RNs, and RTs) at 63.6%. However, both HTx physician and non-physician providers considered EMBs as a necessary part of post-HTx care.

*’But most patients have no issues with biopsy… patients go home usually [in] about 20 minutes… they’ll just be surprised how easy it was.’ – HTx Physician 2*

> *‘I mean, it’s not something that I think people want to do, but they know it’s necessary and part of the maintenance and care of their heart.’ – HTx Non-Physician Provider 8 (APP)*

> *‘Patients have called it pretty uncomfortable in the past but I also think that they find a sense of security with it… it’s like confirmation that, you know, their heart is okay.’ – HTx Non-Physician Provider 9 (HTx RN coordinator)*

While the majority of HTx providers (83.3%) supported a reduction in the frequency of surveillance EMBs, there was also concern of the safety of replacing surveillance EMBs with noninvasive testing, particularly in high risk patients.

> “We still need to see how this protocol rolls out with our higher risk patients… I think we figure that out first, and then maybe it would be a good future goal.” – HTx Non-Physician Provider 11 (HTx RN coordinator)

> *‘[EMB] gives me a sense of comfort in those first few months where [the patients] are really high risk… I feel like we could cut [the EMB frequency] some, but I couldn’t personally see getting rid of them completely.’ – HTx Non-Physician Provider 9 (HTx RN coordinator)*

> *‘Early surveillance [noninvasive testing is] not as reliable. So I think very early on no.’ – HTx Physician 7*

HTx providers (88.9%) also supported many of the suggestions by patients to improve the EMB experience (**Figure 1**). Although HTx physicians agreed on the importance of patient-provider communication, 28.6% expressed reluctance to deliver EMB results electronically, citing risks of heightened patient confusion and anxiety.

> *‘I just think involving the patients. I think that’s why they want to know what the results are even if they don’t understand them. They just want to feel involved… and not just have everybody talking about them like they’re a child… they’re adults and they want to feel empowered.’ – HTx Physician 4*

> *‘I try to check in even in clinics to say, “how are your biopsies going?”… it’s another procedure to us, right, but it’s such a big deal in their life that we kind of miss some opportunities to communicate better to them.’ – HTx Non-Physician Provider 9 (HTx RN coordinator)*

> *‘It’s really a nuanced kind of response looking at the histology… [it might cause] more questions [for] patients.’ – HTx Physician 7*

After making changes suggested by patients and providers (**Figure 1**), we observed a trend in improved patient satisfaction related to EMBs (odds ratio = 1.75; 95% confidence interval, 0.76-4.27; p = 0.155).

## DISCUSSION

To our knowledge, this is the first study evaluating HTx patient and provider perspectives on EMBs and proposed changes to an institutional acute rejection surveillance policy. Our findings identified three key themes: 1) patient anxiety related to EMBs, 2) importance of patient-provider communication, and 3) strong interpersonal trust in providers by HTx patients. Ultimately, HTx patients prioritized the most accurate testing protocol, whether EMB or noninvasive, to safeguard their new heart’s health.

Both thematic analysis and quantitative data confirmed that HTx patients trust providers to select the most effective acute rejection surveillance strategy. However, HTx providers expressed ongoing concerns about the safety and accuracy of early noninvasive testing, particularly in high risk patients.

Previously, others assessed the EMB experience using patient surveys in clinical studies focused primarily on the safety of noninvasive testing.^1,4^ In contrast, we used a mixed-methods approach to examine HTx patient and provider perspectives before any institutional policy changes, as recommended by international guideline standards.^16,17^ Our study revealed that HTx patients offered nuanced priorities for post-HTx surveillance testing, not described before in the literature. In our semi-structured interviews, HTx patients prioritized accurate testing to ensure their heart health over their temporary anxiety from EMBs, which diminished with familiarity. Patients trusted providers to recommend the most accurate surveillance test. Similarly, both HTx physician and non-physician providers emphasized testing accuracy over patient anxiety. These findings align with other medical fields where patients value the clear benefits of accurate, albeit invasive, testing.^18^

HTx patients frequently expressed gratitude for their post-HTx care when reflecting on their EMB experience. Although the recalled pain of EMBs diminished with experience, HTx patients’ gratitude and sense of indebtedness persisted over time. This gratitude was closely tied to strong interpersonal trust in their providers, leading patients to rely on their providers’ judgment when considering changes to institutional policies for acute rejection surveillance strategies. Likewise, patient trust in providers has been recognized as a critical factor in decision-making in other solid organ transplant fields.^19,20^

Our study findings demonstrated that providers sought to decrease the frequency of surveillance EMBs, reflecting the perspectives of the HTx community.^1,21^ However, HTx providers expressed concerns about the safety of early noninvasive surveillance testing for high risk patients. The three RCTs comparing noninvasive testing to surveillance EMBs have only included low risk HTx patients.^2,4,22^ Thus, to convince providers that noninvasive testing can supplant the current standard of surveillance EMBs early post-HTx, more data from RCTs are needed that include high risk patients. Currently, one multicenter RCT (ClinicalTrials.gov NCT06414603) is underway to address this critical knowledge gap.

Even with evolving policies and guidelines on surveillance testing, EMBs will likely stay essential in post-HTx care because of for-cause testing.^15,21^ In our study, both HTx patients and providers offered actionable recommendations for integration into any HTx program (**Figure 1**).

Several limitations should be considered when interpreting the results of this study. First, this was a single center study and may not necessarily represent HTx patient and provider perspectives of other centers. Second, our study focused on proposed policy changes for acute rejection surveillance and thus, our key themes require validation for other aspects of post-HTx care. Third, we acknowledge that most patients did not have complications with their EMBs, which may have impacted their opinions on EMBs. However, previously we found EMB complications to be relatively rare.^15^ In addition, the few patients that did have EMB complications still demonstrated strong interpersonal trust in their providers in our study. Fourth, as our institution has not yet changed our policy for acute rejection surveillance, we were not able to compare results with perspectives after any changes to our institutional policy. However, 9 (32.1%) low, moderate, and high risk patients experienced both EMB and noninvasive surveillance strategies and we found similar results in our thematic analysis. Fifth, volunteer bias is also a potential limitation for our study as patients with positive experiences may have been more inclined to participate. However, we attempted to mitigate this limitation through the use of anonymous online survey responses, allowing for more critical feedback from patients. We found no significant difference in patient age (p = 0.723), sex (p = 0.629), and race/ethnicity (p = 0.570) between the interviewed and non interviewed patients.

## CONCLUSION

We found that the majority of HTx patients prioritized the most accurate acute rejection surveillance strategy and trusted their providers to select it. Our findings offer valuable patient and provider perspectives to inform future clinical practice guidelines.

## Data Availability

The data that support the findings of this study are openly available in Mendeley Data (doi:10.17632/hr6kvn9y4h.1).

https://doi.org/10.17632/hr6kvn9y4h.1

## ACKNOWLEDGEMENTS

The investigators would like to thank all the patients and providers who participated in this study. The authors also acknowledge Michelle H Kim, PharmD, for her review and editing of the final manuscript, and Shelley Orr, RN, Ellen Ashman, NP, Amanda Topik, NP, Amanda Giovanetti, RN, Carmela Zwanziger, RN, Kathleen Cawelti, BSN, RN, Ashley Cardenas, MAS, and Deepa Kurup, RN, MSN, MBA in collecting patient data and distributing the patient satisfaction questionnaires.

## Abbreviations

APP: advanced practice provider
COREQ: Consolidated Criteria for Reporting Qualitative Research
dd-cfDNA: donor-derived cell-free DNA
GEP: gene-expression profiling
HTx: Heart transplant
EMB: endomyocardial biopsy
EMR: electronic medical record
NRC Health: National Research Corporation Health
RCT: randomized controlled trial
RN: registered nurse
RT: radiologic technologist
SRQR: Standards for Reporting Qualitative Research
UCSD: University of California, San Diego

