## Supplementary material for "Perspectives of Heart Transplant Patients and Providers on Acute Rejection Surveillance: A Mixed-Methods Study": supp

**APPENDIX A: 2023 UCSD Post-Heart Transplant Endomyocardial Biopsy Protocol**

| | |
|---|---|
| **Protocol Purpose** | To provide guidelines for Surveillance Endomyocardial Biopsy Schedule for Post-Heart Transplant patients |
| **Definitions** | **High Risk for Rejection:**<br>1. Giant Cell Myocarditis or Cardiac Sarcoidosis.<br>2. Protein-losing Enteropathy with Hypogammaglobulinemia or on Immunoglobulin Treatment.<br>3. Panel of Reactive Antibodies ≥ 40% (can fall under moderate or high risk protocol, case by case discussion).<br>4. Prior Desensitization.<br>5. Positive Crossmatch at Time of Transplant.<br>6. Treated pathologic AMR in the first year.<br>7. ACR > 6 weeks post-transplant.<br>8. Persistent DSA.<br><br>**Moderate Risk for Rejection:**<br>1. ACR in the first 6 weeks post-transplant.<br>2. Isolated historic DSA (without associated acute rejection or graft dysfunction) with repeat testing that demonstrates DSA negative x 2 months.<br>3. Panel of Reactive Antibodies ≥ 40% (can fall under moderate or high risk protocol, case by case discussion).<br><br>**Low Risk for Rejection:**<br>1. Does not meet criteria for moderate or high risk for rejection.<br><br>**Multiorgan Transplant:**<br>1. Heart-Kidney: Same risk stratification as heart only.<br>2. Heart-Lung: Case by case schedule.<br>3. Heart-Liver: Follows low risk heart only protocol unless has high risk features. |
| **EMB Protocol** | **Post-transplant Month 1-3 Schedule:**<br>Right Heart Catheterization and Endomyocardial Biopsy<br>Weeks 2, 4, 6, 8, 10, 12<br><br>**Post-transplant Month 4-6**<br><u>Low risk for rejection:</u><br>Endomyocardial Biopsy<br>Week 16<br>Surveillance with gene-expression profile and dd-cfDNA testing starting Week 16 post-transplant<br><u>Moderate risk for rejection:</u><br>Endomyocardial Biopsy<br>Weeks 16, 20, 24 |

Surveillance with gene-expression profile and dd-cfDNA testing starting Week 24 post-transplant

**High risk for rejection:**

Endomyocardial Biopsy

Weeks 16, 20, 24

**Post-transplant Month 7-10**

Low risk for rejection:

Surveillance with gene-expression profile and dd-cfDNA testing for Weeks 28, 32, 36, and 40 post-transplant.

Moderate risk for rejection:

Surveillance with gene-expression profile and dd-cfDNA testing for Weeks 28, 32, 36, and 40 post-transplant.

High risk for rejection:

Endomyocardial Biopsy

Weeks 28, 32, 36, 40

Surveillance with gene-expression profile and dd-cfDNA testing starting Week 40 post-transplant.

**Post-transplant Indication for Biopsy Outside of Prespecified Testing Time Frame**

Signs or symptoms of rejection or cardiac allograft dysfunction.

dd-cfDNA (AlloSure; CareDx, Brisbane, California, USA) ≥ 0.20% or an absolute increase of dd-cfDNA% of at least 0.05%.[23,24]

Treated rejection outside specified schedules warrants repeat biopsies while weaning steroids.

**Biopsy and HeartCare Schedule**

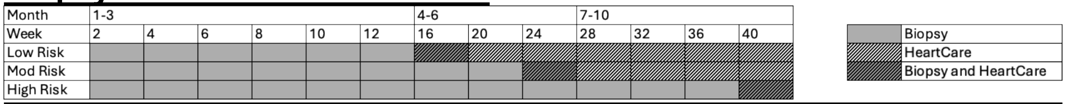


Abbreviations: ACR, acute cellular rejection; AMR, antibody-mediated rejection; dd-cfDNA, donor-derived cell-free DNA test; DSA, donor-specific antibodies; EMB, endomyocardial biopsy.

**APPENDIX B: Semi-Structured Interview Questions for Patients**

1. In your own words, why are endomyocardial biopsies performed as a part of your post-transplant care?
2. How do you feel about endomyocardial biopsies compared to other routine tests you perform as a part of your post-transplant care, such as echocardiograms? What do you think about the frequency of endomyocardial biopsies?
3. What are your thoughts/feelings prior to a scheduled endomyocardial biopsy?
4. What thoughts/feelings do you experience during the endomyocardial biopsy?
5. What are your thoughts/feelings following an endomyocardial biopsy? For instance, how does it affect your activities for the rest of the day and the next day?
6. What are some changes that could be made to improve your experience during an endomyocardial biopsy?
7. Our program is considering further reducing the number of endomyocardial biopsies and replacing them earlier with noninvasive blood tests. What do you think about this proposed change? Do you think this may affect the quality of your care? Do you think this will improve your overall post heart transplant experience?
8. How do you hear about your results for your endomyocardial biopsy? Do you have any suggestions for improving how you get results returned to you?

**APPENDIX C: Semi-Structured Interview Questions for Providers**

1. What words would you use to describe endomyocardial biopsies, both from your perspective and what you have heard from patients?
2. Patients in interviews have suggested topical lidocaine, removal of the sterile face drape, and having the ability to choose their own music during an endomyocardial biopsy. What are your thoughts on these suggestions? Do you think these suggestions could be potentially implemented on a regular basis?
3. Patients in interviews have suggested having the endomyocardial biopsy results available through a patient portal like the rest of their laboratory results with a short interpretation of the results. If results could be displayed with a scripted real-world interpretation, would you think this would be helpful to patients?
4. Patients stated they would feel less anxious and frustrated if they had some way of knowing how much longer they had to wait for their endomyocardial biopsy while in the waiting room. If this could be implemented, would you think this would be helpful to patients?
5. What are you told the purpose of endomyocardial biopsies are for? (question for heart transplant registered nurse coordinators, advanced practice providers, catheterization lab registered nurses, and radiologic technologists)
6. Are you familiar with the potential risks of endomyocardial biopsies? (for all providers)
7. Do you think patients have an understanding of why endomyocardial biopsies are performed and its potential risks?
8. Do you think patients understand what is expected of them during an endomyocardial biopsy?
9. Do you have any suggestions for improving the endomyocardial biopsy experience for patients?
10. There is discussion in the heart transplant field about further reducing the frequency of endomyocardial biopsies and replacing them with noninvasive blood tests, such as donor-derived cell-free DNA testing. What are your thoughts on this topic?

**APPENDIX D: Patient Online Survey Questionnaire**

You are being invited to participate in a research study led by Dr. Paul Kim from UC San Diego Health because you are a heart transplant recipient.

The purpose of this research study is to gain insights and perspectives on the endomyocardial biopsy procedure performed in Heart Transplant patients as well as potential institutional policy changes. Your participation in this research is expected to take approximately 5 to 10 minutes.

Your participation in this study is completely voluntary and you are free to decline to participate in this survey. No identifying information will be collected for this survey. Choosing not to participate or withdrawing will result in no penalty or loss of benefits to which you are entitled. You are free to skip any question that you choose.

All invited participants will be entered into a drawing where they will have a 1 in 10 chance to win $50!

By participating in this research you are indicating that you are at least 18 years old, have read this consent form, and agree to participate in this research study.

This study has been approved by the UCSD Office of IRB Administration (No. 805675) and will be conducted according to accepted and applicable national and international ethics guidelines and principles. If you have questions about your rights as a research participant, or would like to talk to someone outside of the study team about your participation, you may contact the UCSD Office of IRB Administration at or call 85B-246-4777.

---

How much pain do you experience during a biopsy? From 0 (no pain) to 10 (worst pain).

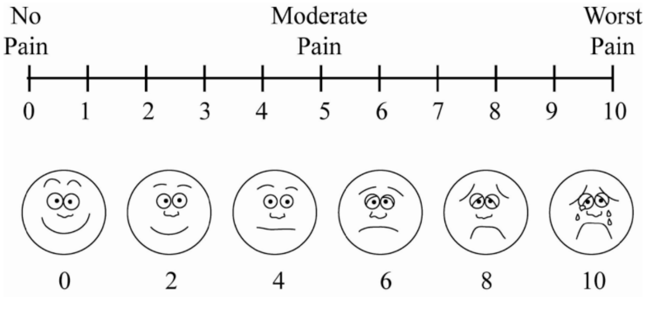


1. An average biopsy?
2. The most painful biopsy you've had?

Please answer how much you agree with the following statements from 1 (strongly disagree) to 5 (strongly agree).

3. Endomyocardial biopsy is an important part of my post-transplant care

4. I initially felt anxious about biopsies, but this improved with experience
5. The physician/staff do a good job communicating with and comforting me during the biopsy
6. My biopsies are typically on time
7. My biopsy results are communicated well to me
8. Following a biopsy, I feel like I can do my normal activities
9. If there was a noninvasive alternative that was as accurate as a biopsy, I would prefer it
10. I would support a change in biopsy frequency based on my team's recommendations
11. The procedure is more comfortable with certain providers compared to others
12. Have you ever had a biopsy where the doctor went through your arm instead of your neck?
13. Which access site did you prefer for your biopsies? (neck vs arm)
14. Is there anything else you'd like to share with us about your experience with endomyocardial biopsy?
